# Longer healthy life, but for how many? Insights on healthy lifespan inequality from the Global Burden of Disease Study

**DOI:** 10.1101/2022.12.06.22283153

**Authors:** Virginia Zarulli, Hal Caswell

**Affiliations:** Interdisciplinary Center on Population Dynamics, University of Southern Denmark; Mathematical Demography and Ecology, Faculty of Science, Institute for Biodiversity and Ecosystem Dynamics, University of Amsterdam

**Keywords:** Healthy life expectancy, health adjusted life expectancy, healthy life expectancy inequality, health variation, global burden of disease

## Abstract

In the last 150 years, in many populations life expectancy has more than doubled, the variation in length of life has decreased, and, as result, more individuals enjoy similarly longer lives (even though with important socio-demographic differences). When it comes to healthy longevity, today more and more people reach older ages in better health than what they used to do only a few decades ago, for many individuals the unhealthy years are getting compressed at the end of life and, overall, healthy life expectancy is increasing globally. But we do not know how many individuals are benefiting from this increase. Indicators of average length of life, such as healthy life expectancy, don’t capture the spread, while similar levels of healthy life expectancy can be achieved by different populations: one where most individuals share a similar number of years in good health, or one where few individuals enjoy high numbers of years in good health compared to many others who don’t. Here we apply demographic techniques for the analysis of variation to the demography of health to study the fundamental question of the distribution of number of healthy years of life among individuals and the relation between healthy lifespan length and healthy lifespan inequality. We use data from the Global Burden of Disease Study, and we produce the first international landscape of healthy lifespan variation over time and by socioeconomic level of the country.

## Introduction

Differences in how many years in good health individuals can expect to live are a fundamental inequality of population health. It has been stated that every inequality depends on being alive or not and that the most fundamental inequality of all is the inequality in the length of life [1]. But being alive *and* healthy is as crucial as being alive, perhaps even more crucial, as most of the relevant dimensions of social and population inequality we can think of, from gender to socioeconomic status, are much more substantiated and affect more the life of the individuals when these are alive and in good health than when they are simply alive. Surviving in good health is the essential basis for acquiring human capital through education, training, experience and social mobility. Healthy years of life are also a crucial part of the dynamics of the life cycle: as long as individuals are healthy they are more likely to be productive, thus being net contributors to the society, as showed by the stronger correlation of the expected number of working years with healthy life expectancy than with (total) life expectancy [2].

Similar levels of healthy life expectancy can be achieved by different distributions of healthy lifespans: one more concentrated, where many individuals share more similar number of years in good health, and one less concentrated, where some individuals enjoy substantially higher numbers of years in good health than other individuals, who suffer from ill health early on. How equally (or unequally) the healthy years of life are distributed between the individuals is one of the fundamental distributional problems of population health. Studying patterns and trends of the distribution of healthy lifespans among the individuals is also important for their crucial socioeconomic implication, for example for retirement and health policies, as highlighted by the analysis of the trends in working life expectancy at age 50 in Europe, which found that this indicator has a higher correlation with healthy life expectancy (based on self-reported health) than with total life expectancy [2]. Shedding light on the patterns of healthy lifespan variation is also relevant for the so called male-female survival paradox, which is the fact that women live longer but have poorer health than men [3, 4]. Recent results suggest that this paradox is a function of indicators of population health and their gender differences [5]. Undoubtedly, healthy lifespan variation is such an indicator and, yet, indicators of healthy lifespan variation are missing from the landscape of demography of health.

Since the concept of healthy life expectancy was introduced by Sanders (6) and later operationalized by Sullivan (7), the increasing interest in healthy longevity has generated a vast literature [8-11] that has focused, however, on expected values and inequalities between groups, disregarding the fundamental distributional question of how the healthy lifespan is distributed between the individuals within the populations. In the last decade or so, researchers working on mortality have recognized the importance of supplementing analyses of average longevity (such as life expectancy) with analyses of variation in lifespan length in the assessment of population health [1], producing a wealth of research [1, 12-24] whose main message is that, as life expectancy has increased, the ages at death have become more similar, even though with some exceptions and differences by gender and social class.

What about the healthy years of life? We know that today people reach older ages in better health than what they used to do only a few decades ago because there has been a general improvement of healthy survival and a postponement of debility [25-28]. But we don’t know what has happened to the distribution of this longer healthy lifespan among individuals: do more individuals enjoy increasingly similar number of years of healthy life, or the distribution has become more unequal?

Some insights come from the literature on the compression of morbidity, which shows a shift to older age of the entry into the morbid (unhealthy) state. These results seem robust despite the numerous differences in the definitions of disability, morbidity and functioning indicators used in the analyses [29, 30], even though there are notable exceptions such as the United States, where an inversion of trend has been reported between 1998 and 2006 [31]. However, the analysis of the compression of morbidity does not equal the analysis of variation in healthy lifespan and is not necessarily informative about the distribution of healthy lifespans within the populations because this is determined by the interaction between the structure of morbidity and of the mortality schedule.

Health adjusted life expectancy (HALE) is a measure of health expectancy that combines both mortality and morbidity: it is the average number of years that a person, of a specified age, can expect to live in “full health.” It considers years lost to mortality and years lived in less than full health due to disease or injury. HALE is used by numerous international organizations, among which the World Health Organization (WHO), and is calculated from the data of the Global Burden of Disease (GBD) Study [32, 33]. We use this measure to produce the first insights on the variation among individuals in the length of healthy life at the global level, using the estimates provided for the 204 countries of the GBD. To our knowledge, this is the first systematic assessment of the trends in healthy lifespan variation. By doing so, we provide the first picture of the international landscape of inequality in healthy longevity among individuals.

The variance among individuals in longevity or healthy longevity calculated from any life table is the result of stochasticity, not of heterogeneity (or, in a strict sense, inequality) among the individuals. The calculations explicitly assume that every individual of a given age is subject to the same demographic rates and thus that differences among individuals in the outcome are the result of chance. Even so, this variation is real and has social and economic consequences. It is also important to note that this variation arises from demographic process and does not represent sampling or estimation errors.

We hypothesize that as healthy life expectancy increased, healthy lifespan variation decreased, following the same negative relation found between life expectancy and lifespan variation. However, it is also possible that the relation between healthy lifespan length and healthy lifespan variation is not the same at all ages as the health systems might be more successful at preventing health deteriorations at some ages rather than at others. For example, for various reasons they might be able or willing to prevent health deteriorations more among children and young adults rather than at older ages.

## Data and methods

We used the estimates of age-specific HALE from the Global Burden of Disease Study [32], a valuable collection of data that provides reliable, comparable healthy and disability information over time, for more than 200 countries around the world from 1990 to 2019.

From the values of HALE, we derived the lifetable that generated those values. Here we call this table “health adjusted life table;” it includes the health-mortality elimination schedule, which we call *q(x)_hale*. This is the probability that an individual in age class *x* leaves the cohort either by death or by losing its “healthy” status. Any column of a life table can be calculated from any other column. The GBD results provide a sequence of healthy life expectancy values *e*(*x*) for ages *x* = 0, 1, 5 and then by 5-year intervals to age 95. Our analysis required us to calculate a sequence of probabilities of death *q*(*x*) compatible with the *e*(*x*) schedule. We used nonlinear least squares to solve for a *q*(*x*) schedule that would minimize the squared deviation between our calculated values and the GBD values for *e*(*x*). The resulting *q*(*x*) schedule was then interpolated to 1-year age intervals with the R-function pclm2D [34]. The survival probabilities that appear in the Markov chain model are given by *p(x)=1-q(x)*. This schedule was used to apply the matrix model (a Markov chain with rewards) developed by Caswell and Zarulli [35] (see the supplementary information and [36] for mathematical details) to estimate the statistics of variation of the healthy life span length. This method considers healthy longevity as a Markov chain with an absorbing state corresponding to death or ill health. An individual surviving from one age to the next gains, as a “reward,” a year of good health. The analysis computes the mean, variance, standard deviation, and other moments of the distribution of health longevity. The mean healthy longevity is exactly the HALE. As a measure of variation, we will focus on the standard deviation of healthy longevity, which we denote as SDHL.

Because of the definition of HALE, *q(x)_hale* represents the age-specific probabilities of exiting the healthy state by mortality or by loss of health due to the acquisition of disability. This is an alternative definition of health from that of Caswell and Zarulli [35], in which age-specific prevalences of disability were used to calculate healthy longevity, implicitly permitting individuals to recover from ill health. The analysis was done for men and women separately.

## Results

From 1990 to 2019, the GBD data shows that HALE at various ages has increased on average in all countries for both men and women. The increment was more substantial at younger ages than at older ages: HALE at birth increased from 59.61 to 64.71 (+5.1 years) for women and from 56.68 to 62.23 for men (+5.55 years), while at age 65 the increase was more modest, from 11.59 to 12.95 (+1.36 years) for women and from 10.01 to 11.39 (+1.38 years) for men.

The picture is more complex when analyzed according to the income level of the countries (according to the World Bank Income Categorization 2019), as showed in fig. 1. HALE at birth has increased, on average, more among low-income countries (+9.49 years and +9.48 years for women and men respectively) than among high-income countries (+3.71 years and +4.79 years for women and men respectively): in the low-income countries HALE at birth is now 57.46 years for women and 55.38 years for men, while in the high-income countries it is 69.67 years and 67.45 years for men and women respectively. On the contrary, HALE at older ages has increased more among high income countries than among lower-income countries. For example, HALE at age 50 for women has increased by 2.65 years in high income countries and 1.86 years among low-income countries. The values for men are 3.2 years and 1.93 years respectively. HALE at age 15, instead, has increased more among women in low-income countries than in high income countries (+3.7 years vs +3.1 years) but more for men in high income countries than in low-income countries (+3.55 vs +4.06 years).

**Fig. 1.**
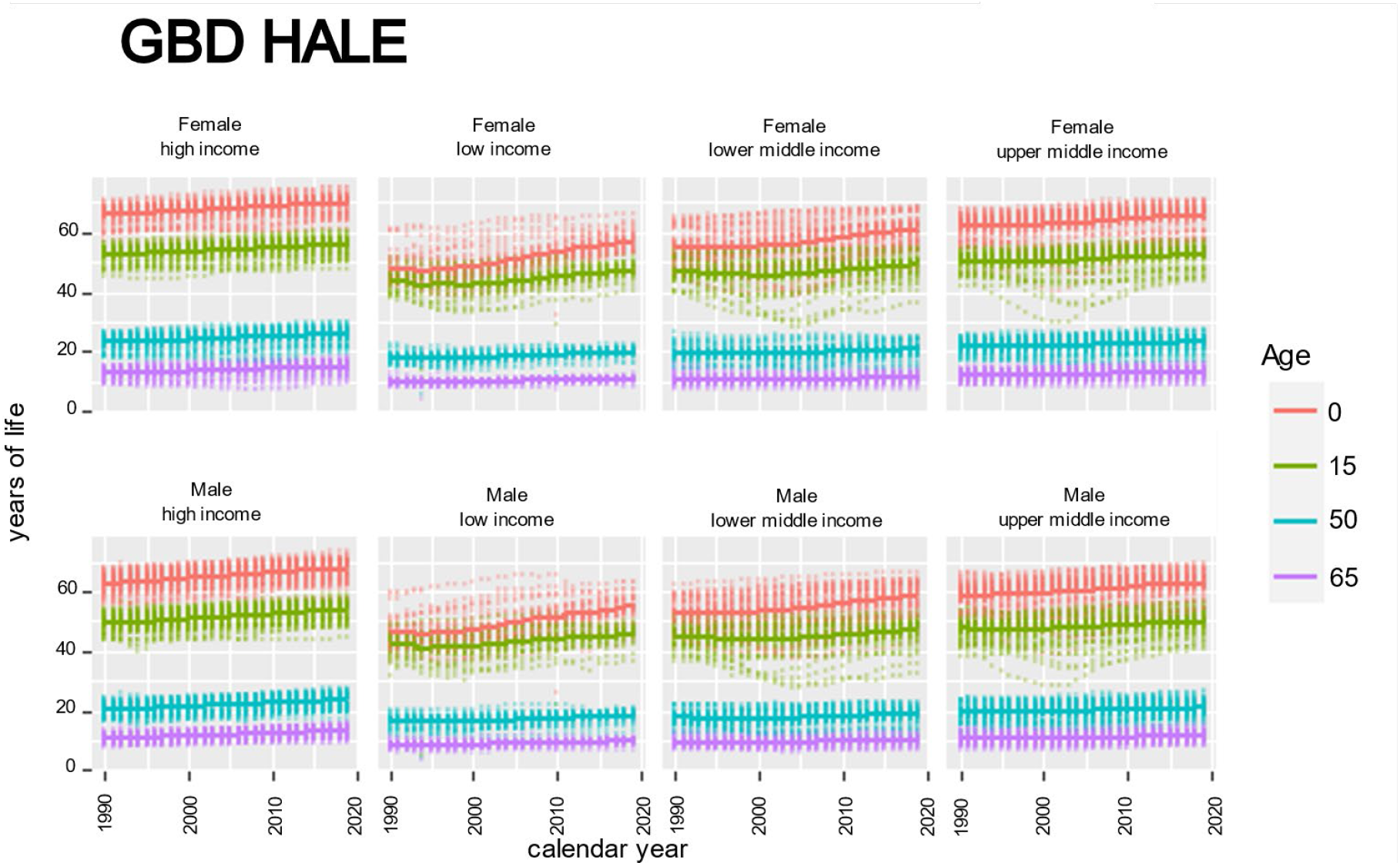
HALE values by sex and country income level over time at age 0, 15, 50 and 65. All countries (faded points) and average value (solid lines). Source: Global Burden of Disease

The analysis of variation among individuals in healthy longevity shows an inverse trend: on average, SDHL has decreased at younger ages similarly for both men and women: at age 0 the reduction was of 2.13 years for women and 2.16 years for men and at age 15 it was of 0.74 years and 0.68 years. On the contrary, inequality in HALE among individuals at older ages has slightly increased, and slightly more for men than for women: for example, at age 65 the increased was of 0.23 years for men and 0.16 years for women.

When analyzing the trends by country income groups (fig. 2), it is striking how the reduction of SDHL at birth (and to a lesser extend also at age 15) has been much more substantial among low-income countries than among high income countries, with 3.5 to 5 times bigger reductions: -3.33 years vs -0.67 years for women and -3.03 years vs -0.88 years for men. At older ages, instead, variation has increased, and it has done so more among high income than among low-income countries, for both men and women: for example, at age 60, in high income countries SDHL grew by 0.23 years for women and by 0.38 for men, while in low-income countries the increment was of 0.16 and 0.19 years for women and men respectively.

**Fig. 2.**
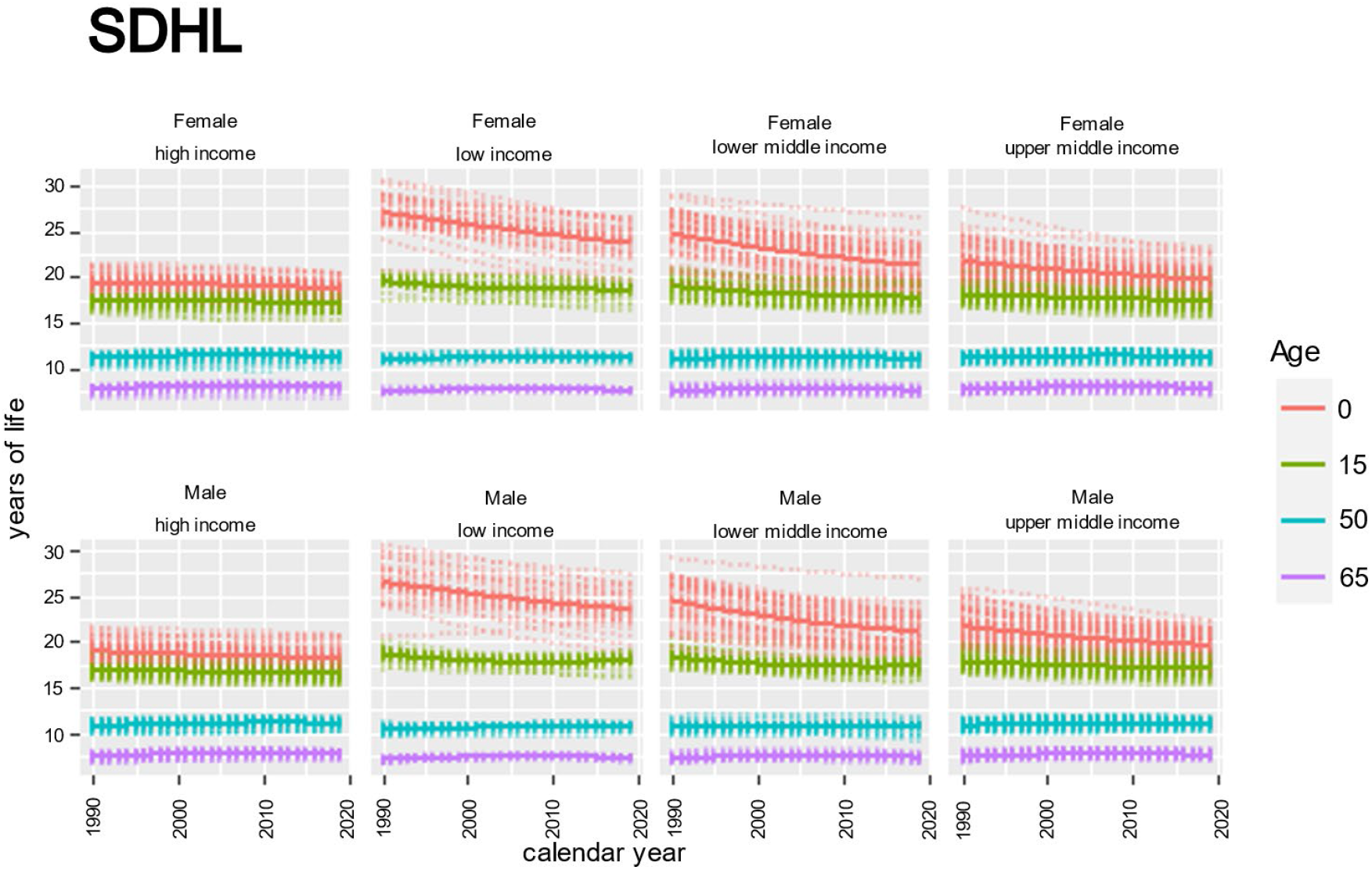
Variation in healthy longevity (measured as SDHL) by sex and country income level over time at age 0, 15, 50 and 65. All countries (faded points) and average value (solid lines). Source: authors’ calculations

Moving beyond the analysis of average trends over time allows to uncover a much stronger and regular relation, different by age, between healthy lifespan length and variation in healthy lifespan. Fig. 3 reports the pairwise relation, for each country in each year, between HALE and SDHL. At birth (and less strongly at age 15), the relation is negative and all the countries, independently of their income level, have decreased the level of variation while increasing healthy life expectancy. The tendency has been more pronounced for low and lower middle-income countries than for upper middle- and high-income countries.

**Fig. 3.**
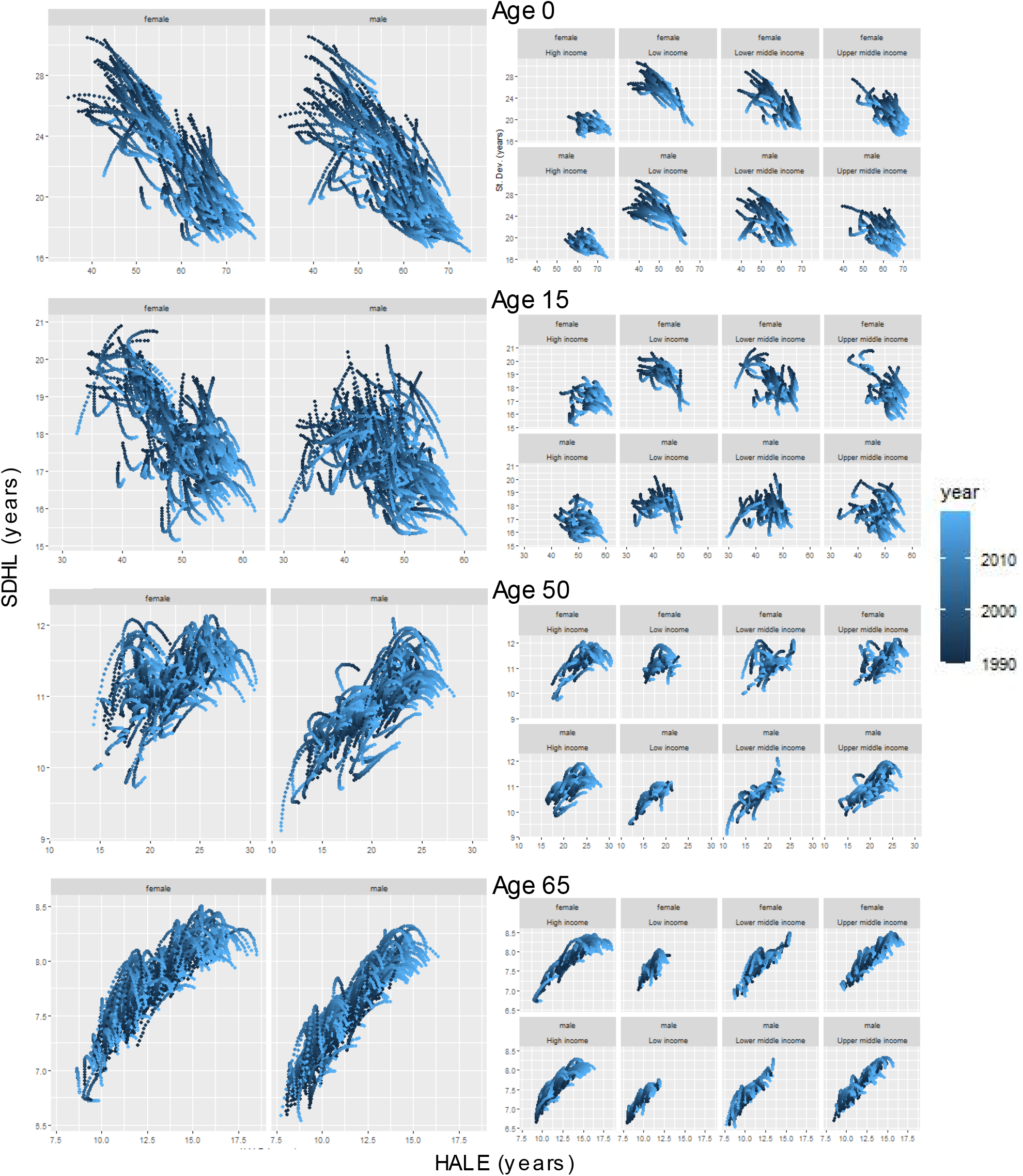
Relation between healthy lifespan length (HALE) and healthy lifespan variation (STHL) at different ages, for men and women and different levels of country income, from 1990 to 2019.

At adult ages the situation changes and the relation between healthy lifespan length and healthy lifespan variation becomes positive. Also, this relation appears valid in all the countries and to be strengthening with age. However, at adult ages the positive relation is more pronounced among upper middle- and high-income countries. Most notably, these countries have increased both the mean and the standard deviation of healthy life span more than the lower middle- and low-income countries.

Women tend to have longer healthy lifespan and higher healthy lifespan variation than men, as indicated by the male to female ratio of both indicators, HALE and SDHL, smaller than 1. Figure 4 shows the density function of these ratios for age 0 and figure 5 for age 65. Over time the ratios are consistently below 1 for most of the countries. It is worth noticing that the cases of longer healthy lifespan for men compared to women (and of higher variation) are mostly located in low-income countries compared to high-income ones, both at age 0 and at age 65.

**Fig. 4.**
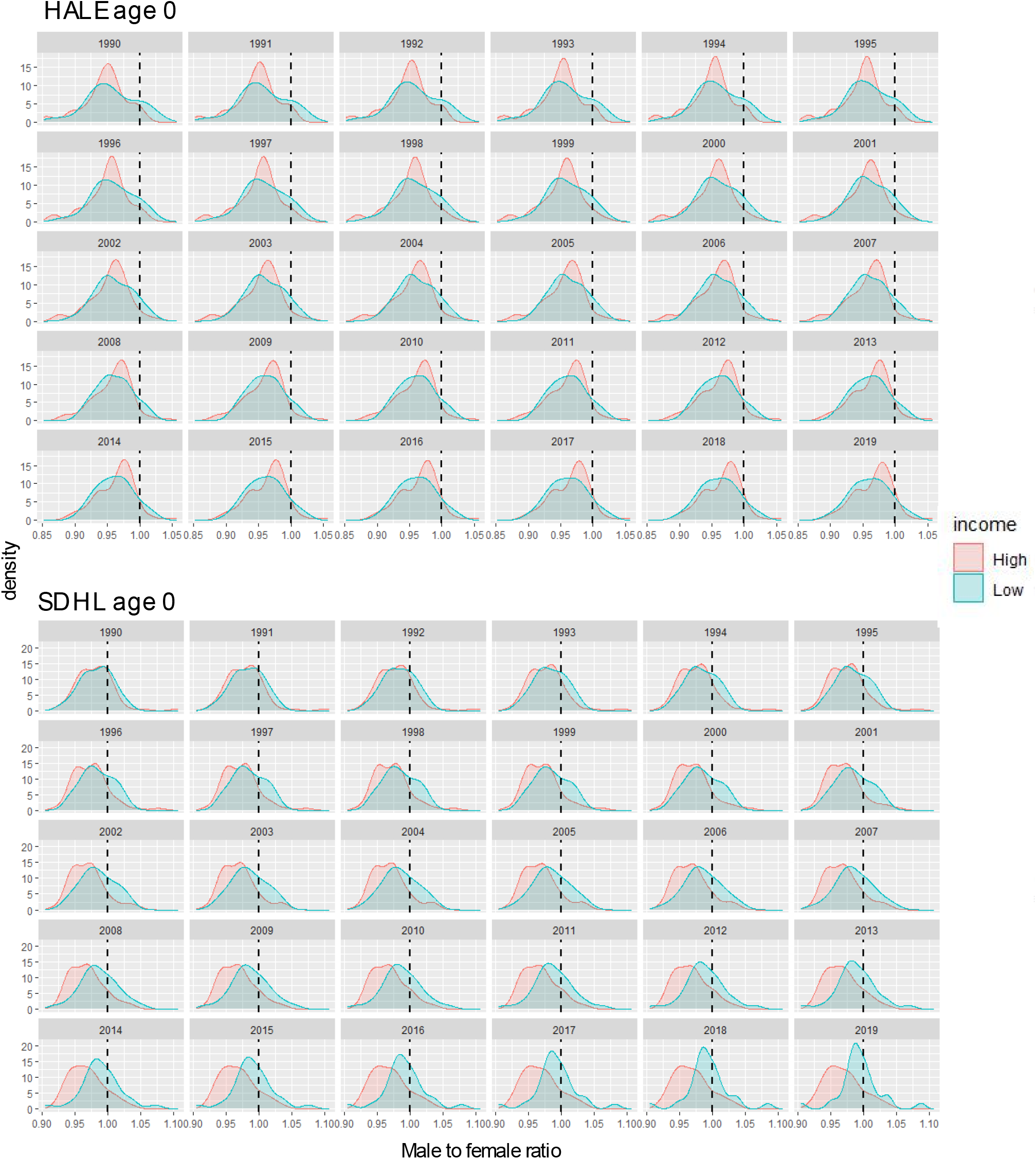
Male to female ratio of healthy lifespan indicators at birth for high-income and low -income countries, from 1990 to 2019. Healthy lifespan length, as measured by HALE (upper panel); healthy lifespan variation, as measured by SDHL (lower panel).

**Fig. 5.**
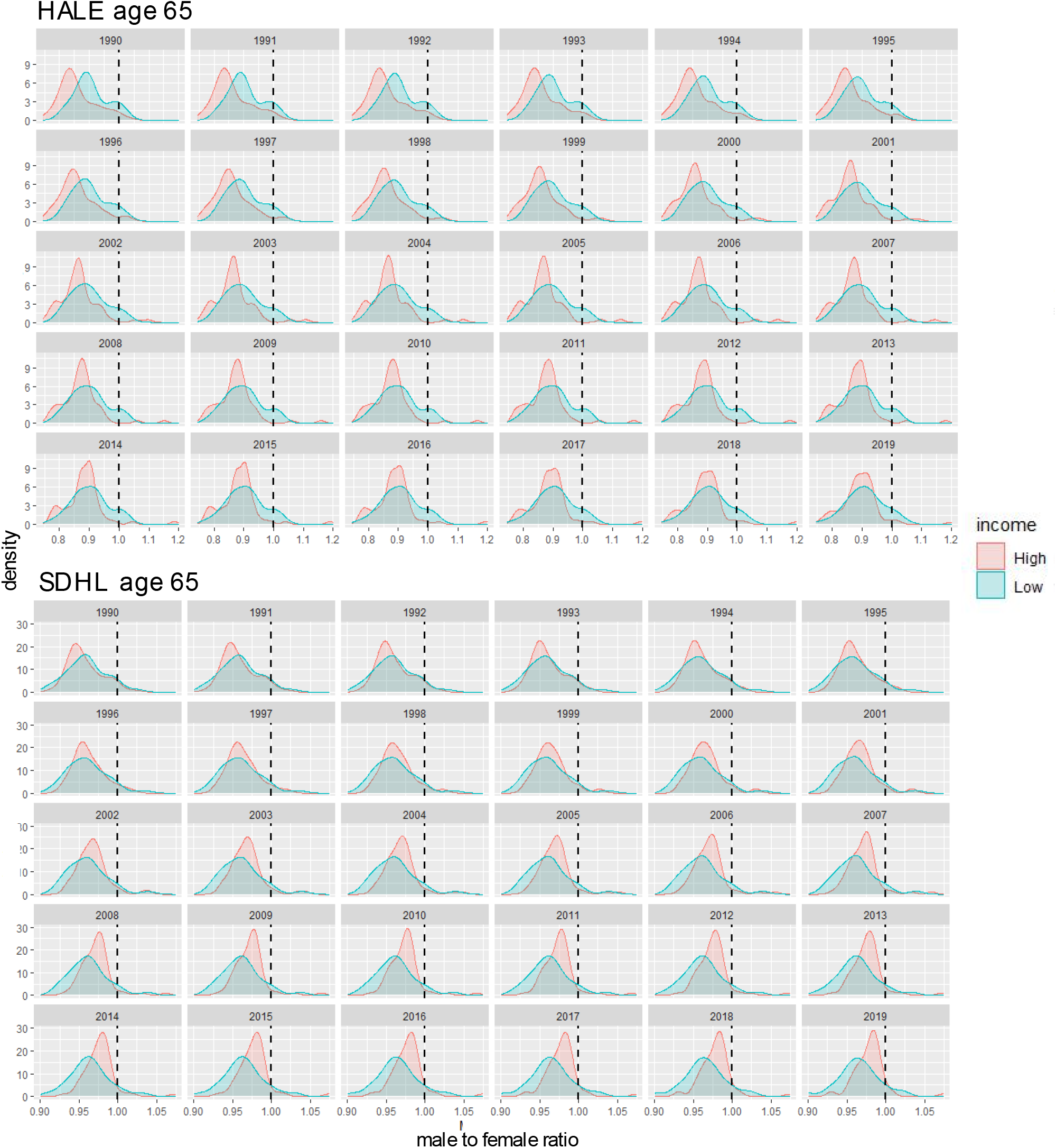
Male to female ratio of healthy lifespan indicators at age 65 for high-income and low - income countries, from 1990 to 2019. Healthy lifespan length, as measured by HALE (upper panel); healthy lifespan variation, as measured by SDHL (lower panel).

From 1990 to 2019, among high-income countries, male and female healthy life expectancies have become more similar (more at birth than at age 65), as showed by density of the ratio of male to female HALE, that has become more concentrated and whose peak has gradually shifted toward 1 – figure 4 and 5, upper panels. A similar trend, but much less pronounced, can be spotted also among low-income countries. Nevertheless, women still benefit from longer healthy life expectancy than men. This difference is particularly pronounced at age 65.

Men and women had, instead, remarkably divergent trends of variation at age 0 in healthy year of life lived among high- and low-income countries - figure 4 and 5, lower panels. While the ratio in most cases remains solidly below 1, indicating a higher female variation in terms of healthy years of life, the densities for the high- and low-income groups, practically overlapping in 1990, have diverged substantially over time until 2019. At age 0, changes over time have made the two groups of countries diverge: in low-income countries the male to female ratio moved towards 1 and in high-income countries it moved further from 1. The male to female difference in variation of healthy living at age 65, on the contrary, underwent changes over time almost exclusively among high income countries, where the ratio became sharply concentrated around values closer to 1.

## Discussion

In the last decades healthy life expectancy has increased all around the world but we do not know how many individuals are benefiting from this increase because measures of variation of healthy longevity have not been systematically analyzed. This study of healthy lifespan variation among individuals will fil a current gap in literature in the field of population science and will prove necessary, if longevity is to be socially and economically sustainable in societies where the number of individuals reaching old ages is growing, because uncertainty in lifespan is costly, both economically and medically [37].

We have showed that the relation between healthy lifespan and healthy lifespan variation differs with age. It is negative at young ages and positive ad adult, old ages. As healthy life span at birth and at younger age increased, the variation in healthy lifespan has decreased. As healthy life expectancy at adult and older ages increased, instead, the variation increased. For reduction in variation to happen, it is necessary that the interaction between the age structure of survival and the age-structure of morbidity produces health improvements at ages below specific threshold ages, below which the improvements are translated into a reduction of variation [12, 23]. On the contrary, improvements that happen at ages above the threshold ages translated into an increase of variation. Our results of a decreasing variation when considering the whole age-range or almost (from birth or form age 15), to increasing variation when focusing only on older ages (from 50 or from 65), indicates that improvements in healthy survival are happening across all the age range. To assess to what extent and with which age-specific differences requires the analysis of the age specific contributions to the relation between healthy life expectancy and healthy lifespan variation. Such analysis goes beyond the scope of this paper, even though it is a relevant and complex topic worth of investigation in a future study.

The most pronounced increase in healthy life expectancy at birth happened among the lower- and lower-middle income countries, which also experienced the sharpest reduction in healthy lifespan variation. Consequently, these countries have reduced the gap with the high- and upper middle-income countries in terms of both average length of healthy life and individual variation around it. This is encouraging and means that from low-to high-income countries, the span of healthy life at birth is getting longer and more and more individuals are benefiting from this prolongment (as indicated by the reduction of the variation).

At older ages, instead, the increase in healthy life expectancy has been more substantial among the high- and upper middle-income countries, as well as the increase of the level of variation. This is a double-sided phenomenon: on the one side, these countries are experiencing a sizeable improvement in healthy survival also at older ages, on the other side, these improvements are more and more unequally distributed among the individuals. Whether this is only due to the formal relation between the improvements in the age-specific healthy survival rates and the dynamics of healthy lifespan-healthy lifespan variation [12, 23], or also to other factors, for example social dynamics, is an important open question awaiting for more research.

The high-low-income countries patterns that we have highlighted partly resemble the phases of the life expectancy revolution [38] and the epidemiologic transition [39], which happened first in a group of leading countries, and then gradually spread to the others. The striking leap forward in survival rates and the shift to a disease pattern dominated by degenerative, chronic, and man-made diseases (which tend to affect older ages), have more than doubled the life expectancy and reduced the individual variation and uncertainty around the age at death. The major increases in survival start with dramatic improvements at birth and then gradually shifts to older ages. By the time the forerunners reach the stage where improvements are still possible (almost) only at old ages, the late comers are tackling the initial stage of the phase and experience significant survival improvements at young ages.

The dynamics that we have described so far are not the same for men and women and the gender trends appear different depending on the income level of the country. We have found that women tend to have longer healthy lifespan and higher healthy lifespan variation than men. We also have found that over time women tend to have both a longer mean healthy lifespan and higher variation in healthy lifespan than men, with fewer cases where men have longer healthy lifespan and higher variation compared to women. However, a breakdown of the data by income level of the country reveals, moreover, that these cases are located more in low-income countries than in high-income ones, both at when looking at the indicators of healthy longevity at age 0 and at age 65. Overall, men and women in both groups of countries have experienced an increase of healthy life expectancy, but this was more pronounced among men and in high-income countries. Men and women had, instead, divergent trends of variation. When considered at age 0, in low-income countries, men and women have gradually moved towards similar level of healthy lifespan variation, while in high-income countries they have become more and more different, with women experiencing higher levels of variation than men in most cases. When considered at age 65, sizeable changes (towards more similar level of healthy lifespan variation between the two sexes) were detected only among high-income countries.

## Data Availability

All data produced in the present study are available upon reasonable request to the authors

## Authors contributions

*CRediT roles: Conceptualization; Data curation; Formal analysis; Funding acquisition; Investigation; Methodology; Project administration; Resources; Software; Supervision; Validation; Visualization; Roles/Writing - original draft; Writing - review & editing*

Virginia Zarulli: Conceptualization, Data Curation, Formal Analysis, Investigation, Methodology, Visualization, Writing - original draft, writing - review and editing.

Hal Caswell: Methodology, Writing - review and editing.

## Ethics approval

This study did not require ethical approval, as it is an observational study that uses aggregated, population level, publicly available data.

## Supplmentary information

### Calculating survival probability from HALE

Any column of a life table can be calculated from any other column. The GBD results provide a sequence of healthy life expectancy values *e*(*x*) for ages *x* = 0, 1, 5 and then by 5-year intervals to age 95. Our analysis required us to calculate a sequence of probabilities of death (*q*(*x*) compatible with the *e*(*x*) schdule. We used nonlinear least squares to solve for a *q*(*x*) schedule that would minimize the squared deviation between our calculated values and the GBD values for *e*(*x*). We constrained the final *q*(*x*) value to be 1.0. The method also assumes the *a*(*x*) values are known. In a traditional lifetable setting, the *a*(*x*) values represent the number of person-years lived in the interval x by those who exit the population in that interval. This is usually approximated by half of the interval (this means that, on average, the individuals die halfway through the interval). For our estimation, in the absence of specific literature on *a*(*x*) values for a health adjusted life table, we have adopted a conservative approach by assuming that those who leave the healthy state (by death or by acquisition of disability) do so, on average, half-way through the interval. Therefore, we used *a*(*x*) values equal to half of the length of the age interval.

The resulting *q*(*x*) schedule was then interpolated to 1-year age intervals using the R-function pclm2D [2]. The survival probabilities that appear in the Markov chain model are given by *p*(*x*) = 1 *− q*(*x*).

### Markov chains with rewards

We calculated the mean, variance, and standard deviation of healthy longevity using Markov chains with rewards [1] (see [3] for mathematical details). The Markov chain transition matrix describing the life of an individual is given by (shown here for 3 age classes)

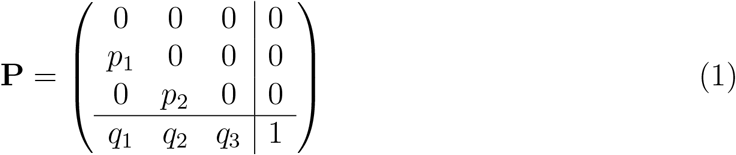

The subdiagonal entries are age-specific survival probabilities and the last row contains age-specific probabilities of death or loss of health.

Lifetime accumulation of healthy life is treated as a “reward” obtained for every transition in the individual’s life. Surviving a year in good health yields a reward of one year. Death or loss of health yields *a*_*x*_ partial years of healthy life. The matrix of the first moments of rewards is

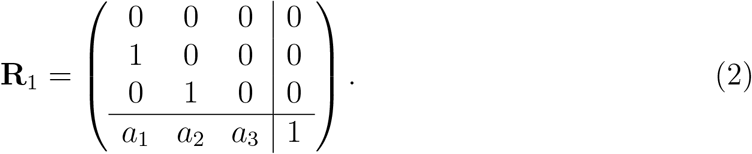

Because the rewards are fixed, the second moments are

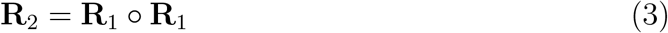

where ◦ denotes the element-by-element product.

The vectors of the first moments and second moments of healthy longevity are calculated using equations (8) and (9) of [1]. The first moments are the mean healthy longevity (HALE). The variance and the standard deviation (SDHL) are calculated from the first and second moments using equations (14) and (15) of [1].

A comparison of our estimates of HALE with those reported by GBD revealed an excellent correspondence until age 80 (fig. 1).

**Figure 1:**
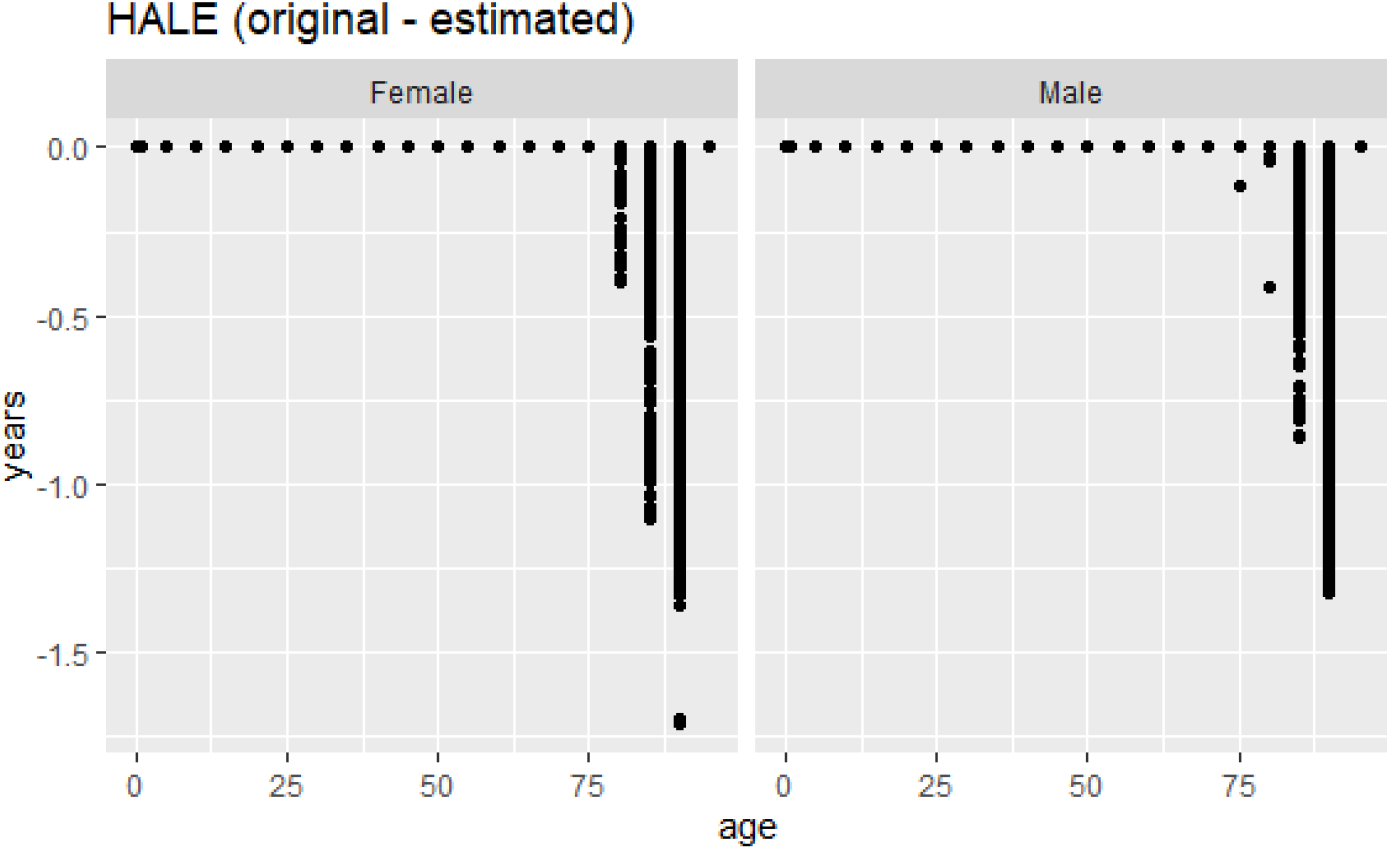
Estimated HALE from authors’ calculation minus HALE reported by Global Burden of Disease (GBD)

